# Errorless learning of a video call in Alzheimer Disease: verbal–motor error dissociation as a potential marker of procedural learning (a pilot study)

**DOI:** 10.64898/2026.09.08.26362385

**Authors:** Caroline Vincenti

**Author notes:** Corresponding author: Caroline Vincenti, Department of Philosophy, Université de Montréal, Montréal, Québec, Canada.

## Abstract

**Background:** Dementia is rising worldwide, and in Alzheimer Disease (AD) the loss of communication adds to the burden. In AD the explicit, error correcting memory fails early, whereas procedural and implicit memory is relatively preserved, which is the basis of errorless learning.

**Objective:** To test whether people with AD can learn an everyday communicative task, placing a video call, through errorless learning, and to characterise the learning curve and the pattern of verbal and motor errors.

**Methods:** Twenty patients with AD learned a three step video call on a touchscreen through errorless learning, over two sessions of 15 minutes with three trials each, separated by a 15 minute interval. Execution time was recorded for each trial. Verbal errors, a correct gesture with an incorrect verbalisation, and motor errors, a wrong, approximate, or absent gesture, were scored for each step. Means are reported with 95% confidence intervals.

**Results:** Mean execution time fell from 41.7 to 23.3 seconds and levelled off over the last two trials of the second session. Verbal and motor errors followed significantly different courses across steps: verbal errors were present from the first trial and fell close to zero by the last step of the second session, whereas motor errors increased.

**Conclusion:** People with AD acquired the procedure, and the dissociation between verbal and motor errors is consistent with a dissociation between procedural performance and explicit verbal report during acquisition. These preliminary findings support the use of preserved procedural memory in rehabilitation.

## 1. Introduction

Dementia is rising fast, and in Alzheimer Disease (AD) communication loss feeds a vicious circle of isolation and decline (GBD 2019 Dementia Forecasting Collaborators, 2022; Klimova et al., 2015). This raises a core clinical problem, because the explicit, error correcting memory that conventional rehabilitation depends on is the first system to fail (Baddeley & Wilson, 1994).

We therefore turn to errorless learning, a rehabilitation method that prevents errors during acquisition and leans on relatively preserved implicit memory, and its procedural component in particular, rather than on the damaged explicit memory (Baddeley & Wilson, 1994; Beaunieux et al., 2006). Errorless learning is already used in neurological rehabilitation, in dementia (de Werd et al., 2013) and in communication disorders such as aphasia (Fillingham et al., 2006; Nunn et al., 2023), and people with mild dementia can relearn tasks involving technology (Kerkhof et al., 2022). If it works here, people with AD can relearn a concrete communicative act, placing a video call, keeping a channel of communication open (Joubert et al., 2022).

Our aim is therefore analytic rather than merely feasibility-based. Scoring verbal errors, defined as a correct gesture paired with an incorrect verbalisation, apart from motor errors lets us read this dissociation as a marker of the parallel engagement of implicit and declarative retrieval across acquisition (Cohen & Squire, 1980), which refines Ackerman’s (1988) account of skill acquisition. For the rehabilitation team, and the speech and language therapist in particular, it offers a concrete way to use preserved procedural memory to help maintain communication, while these findings remain preliminary (Kessels & Olde Hensken, 2009).

## 2. Materials and methods

### 2.1. Participants

Twenty patients with Alzheimer Disease took part in the study and completed both sessions. They were recruited at a municipal day care centre for older adults in Nice, France. Data were collected in 2012. The inclusion criterion was a Mini-Mental State Examination (MMSE) score of 10 or above, and none of the patients used a computer at home. The sample comprised 17 women and 3 men. Mean age was 84.3 years (95% CI 82.0 to 86.5), and the mean MMSE was 20.4 (95% CI 18.2 to 22.6), consistent with mild to moderate Alzheimer Disease. As a pilot study, sample size was based on the number of eligible patients attending the day care centre; no a-priori power calculation was performed.

### 2.2. Materials and measures

The task was to place a video call on a computer. It involved three steps: opening the application icon, selecting the contact, and pressing call. The screen was used as a touchscreen, since the mouse proved impractical, and patients responded with their index finger. Visual cues supported each step. The rest of the screen was kept black to remove distractors, the target icon and a red guiding arrow were enlarged, and windows opened with a single click to avoid double click difficulties.

Two outcomes were recorded. Execution time was measured for each trial. Errors were scored at each step and kept in two separate categories. A verbal error was a correct gesture accompanied by an incorrect verbalisation, for example saying that one should go to the left while correctly clicking on the right. These verbal errors are propositional misstatements about the direction of the ongoing action and are distinct from the lexical and semantic retrieval failures that also characterise AD. A motor error was a wrong, approximate, or absent gesture.

### 2.3. Procedure

The protocol had two phases. In the explanatory phase, the task was described and then demonstrated with errorless learning. The experimenter pointed to the path while describing it aloud, combining a visual and an auditory cue to support encoding, and the patient then carried out the path. In the learning phase, the patient practised over two 15-minute sessions separated by a 15-minute interval of usual activity, with three trials per session, each requiring the three steps.

### 2.4. Statistical analysis

Mean execution time was computed for each trial and each session, with 95% confidence intervals, and we noted the trial at which the decrease levelled off, that is the preliminary plateau. Error counts per step were analysed with a Poisson generalised linear mixed model, with error type, step, session, and the error type by step interaction as fixed effects, and a random intercept for each participant. The interaction, tested by a likelihood ratio test, is the test of the dissociation, and was confirmed by a robust population averaged model and a patient level signed rank test. All 20 participants completed both sessions and no data were missing.

## 3. Results

Execution time decreased across the six trials, from 41.7 seconds (95% CI 26.3 to 57.0) on the first trial of session 1 to 23.3 seconds (95% CI 15.6 to 31.0) on the last, steep at first and then levelling off. The last two trials of session 2 were almost identical, at 24.5 and 23.3 seconds, suggesting a preliminary plateau (Figure 1).

**Figure 1.**
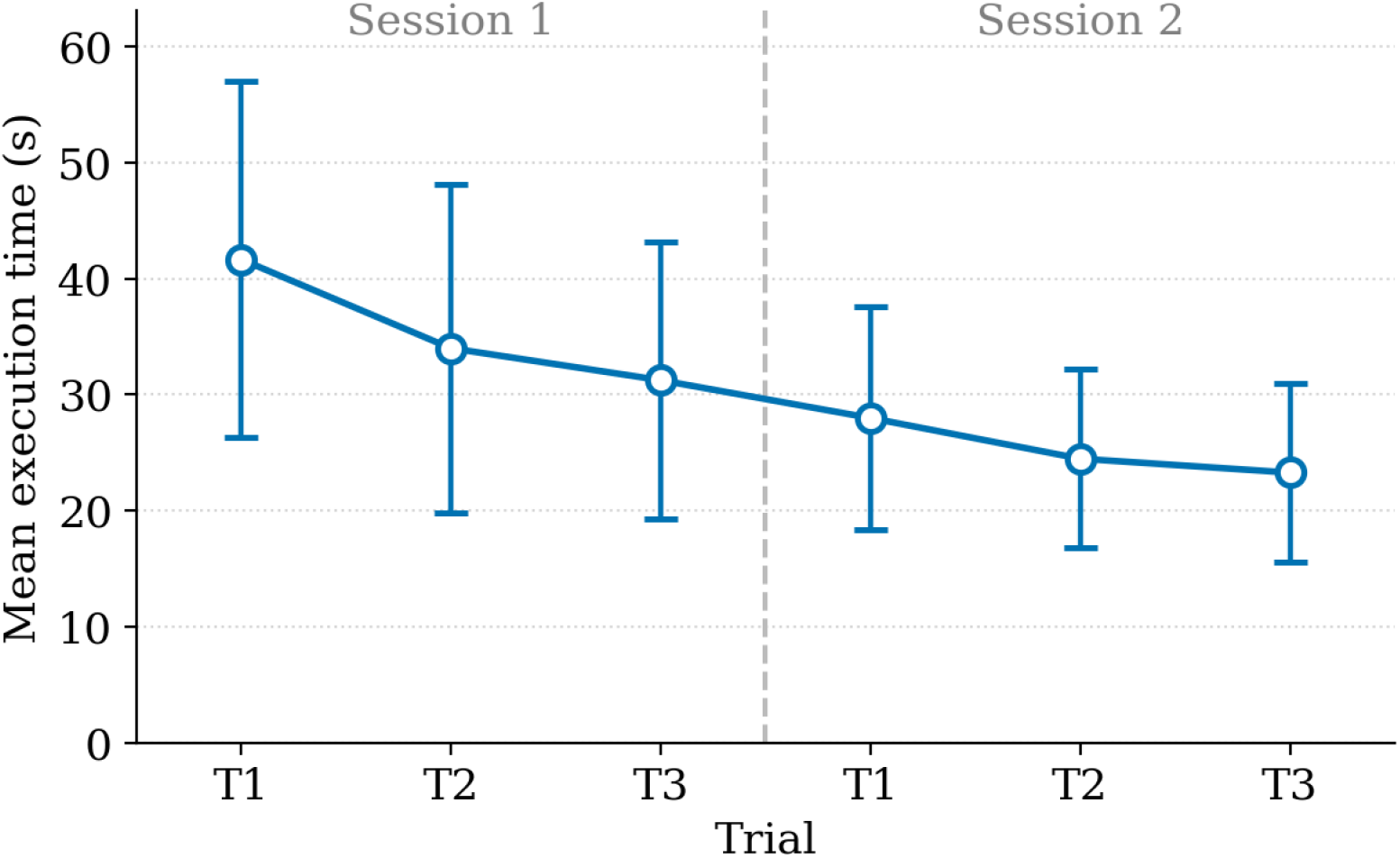
Mean execution time per trial across the two sessions, with 95% confidence intervals. The dashed line separates session 1 from session 2. The decrease is steep at first and then levels off over the last two trials.

Error counts were low throughout, and the two error types followed different courses. Verbal errors were present from the first trial. Across the three steps the verbal error curve was concave, highest at step 2 and lowest at step 3, and verbal errors were close to zero by the third step of session 2. Motor errors, by contrast, rose across the three steps, by about fourfold from the first to the third step (Figure 2). This difference between the two step profiles was significant (error type by step interaction, likelihood ratio chi square 18.3, two degrees of freedom, p = 0.0001), and was confirmed by a population averaged model (p = 0.014) and a patient level signed rank test (p = 0.001).

**Figure 2.**
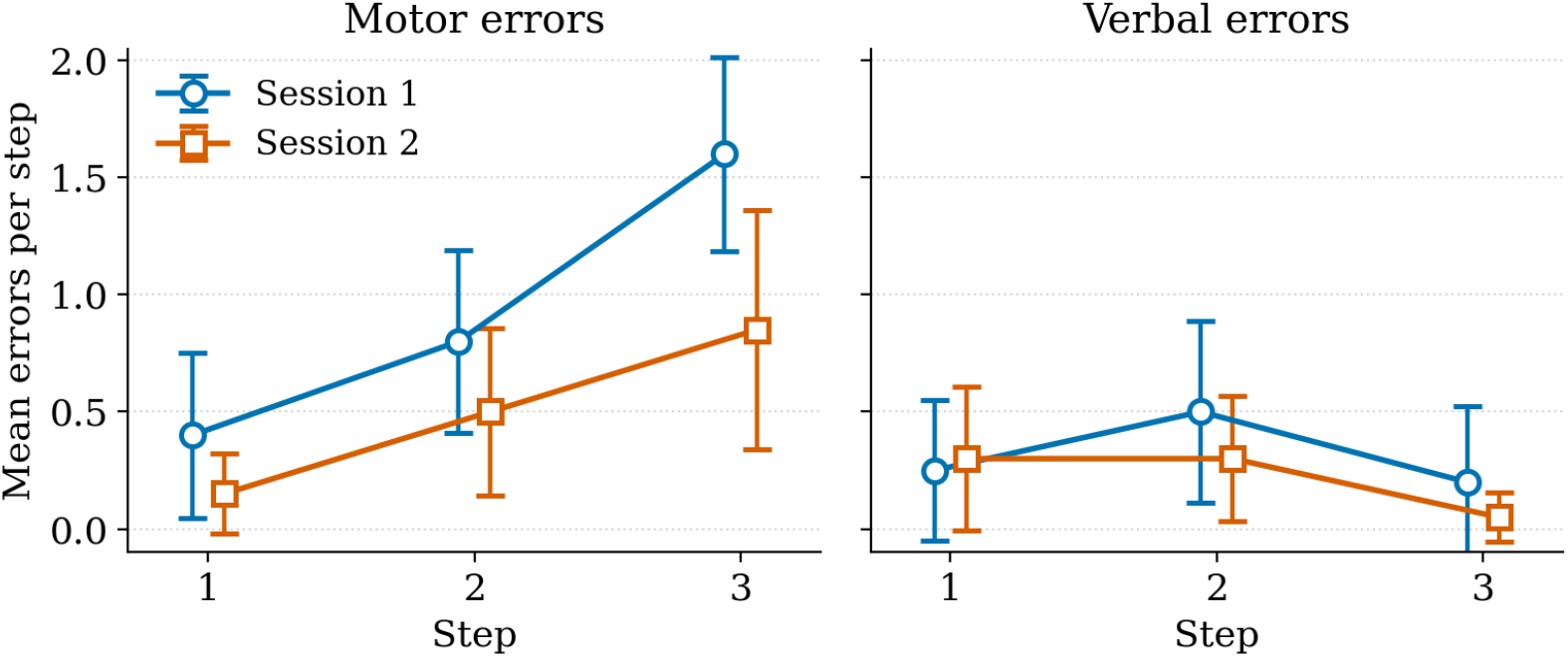
Mean motor errors (left) and verbal errors (right) per step, for session 1 and session 2, with 95% confidence intervals. Motor errors increase across steps, whereas verbal errors are concave and fall close to zero by the last step of session 2.

## 4. Discussion

This pilot study shows that people with Alzheimer Disease can acquire a concrete, everyday communicative procedure, placing a video call, through errorless learning. That such procedures can be learned is consistent with earlier controlled work in dementia (Kessels & Olde Hensken, 2009; de Werd et al., 2013), so the feasibility is not in itself new. Our contribution lies in the error analysis.

The two error types behaved differently. A verbal error marks a moment when the patient performs the procedure correctly while verbalising it incorrectly, which we read as a marker of implicit retrieval. Verbal errors were present from the very first trial and fell close to zero by the last step of the second session (Figure 2). This suggests that implicit memory may contribute from early in skill acquisition, and not only in its final autonomous phase, which refines Ackerman’s (1988) account.

Two qualifications matter. First, the verbal errors scored here are misstatements about the direction of the ongoing action, not lexical retrieval failures, and so are not reducible to the semantic and word finding deficits also seen in AD. Second, because errorless learning models the correct gesture at the outset, a correct gesture on the very first trial may reflect model following rather than implicit retrieval per se. The cleaner signal is the maintenance of the correct gesture across subsequent trials, when no demonstration is given and verbalisation remains incorrect.

Motor errors followed the opposite course and increased across the three steps. This is at least partly explained by the application interface, which became harder to read from the second step on, so the increase reflects display and task constraints rather than a failure of procedural learning. That the fall of verbal errors reflects acquisition rather than the intrinsic difficulty of a given step is supported by the across-session comparison: at the second and third steps, verbal errors fell from the first to the second session, with the step held constant.

### 4.1. Limitations

Several limitations temper these conclusions. There was no control group, so we make no claim of superiority over trial and error learning (Voigt-Radloff et al., 2017). The central finding, however, is a within-participant dissociation in which each patient serves as their own reference, so it does not depend on a control group. Error counts were low, so the model should be read as exploratory, even though three analyses agreed. Errors were scored by the experimenter during sessions, without independent blinded coding or inter-rater reliability assessment, which limits the robustness of the error classification. The motor increase is partly an interface effect, so the divergence should not be read as purely cognitive on the motor side. The 2012 software has since been discontinued, though the procedure transfers to current video calling tools. Finally, the durability of the learning was not assessed, and the directive nature of errorless learning limits the patient’s active engagement.

## 5. Conclusion

A short procedural communicative task can be relearned in Alzheimer Disease through errorless learning, and the dissociation between verbal and motor errors is consistent with a dissociation between procedural performance and explicit verbal report during acquisition. Clinically, this supports leaning on preserved procedural memory to help maintain communication. The findings are preliminary and call for a larger controlled study, a test of long term retention, and an evaluation of transfer to the home setting.

## Statements and Declarations

### Ethics approval and consent to participate

This non-interventional study was carried out in France, in 2012, with patients attending a municipal day care centre. Under the loi Huriet-Sérusclat of 1988, which was the applicable framework at the time, non-interventional research of this kind did not require review by a Comité de Protection des Personnes. Mandatory ethics committee review for all categories of non-interventional research was introduced only later, by the loi Jardé, in force from November 2016. The study followed the principles of the Declaration of Helsinki, and all data were fully anonymised and analysed in aggregate, so that no individual participant can be identified.

Ethics approval for this secondary analysis was granted by the Research Ethics Committee for Society and Culture (CERSC) of Université de Montréal (reference number 2026-8684, 29 June 2026).

The original 2012 study was non-interventional and did not require written informed consent under the French research framework applicable at the time (loi Huriet-Sérusclat of 1988). All data used in the present secondary analysis are fully anonymised and analysed in aggregate, so no individual participant can be identified.

## Data availability

The data that support the findings of this study are available from the author upon reasonable request.

